# Quantifying the impacts of human mobility restriction on the spread of COVID-19: an empirical analysis from 344 cities of China

**DOI:** 10.1101/2020.07.13.20148668

**Authors:** Jing Tan, Yiquan Xiong, Shaoyang Zhao, Chunrong Liu, Shiyao Huang, Xin Lu, Lehana Thabane, Feng Xie, Xin Sun, Weimin Li

## Abstract

**Objective:** Since the outbreak of novel coronavirus pneumonia (COVID-19), human mobility restriction measures have raised controversies, partly due to inconsistent findings. Empirical study is urgently needed to reliably assess the causal effects of mobility restriction.

**Methods:** Our study applied the difference-in-difference (DID) model to assess declines of population mobility at the city level, and used the log-log regression model to examine the effects of population mobility declines on the disease spread measured by cumulative or new cases of COVID-19 over time, after adjusting for confounders.

**Results:** The DID model showed that a continual expansion of the relative declines over time in 2020. After four weeks, population mobility declined by 54.81% (interquartile ranges, −65.50% to −43.56%). The accrued population mobility declines were associated with significant reduction of cumulative COVID-19 cases throughout six weeks (i.e., 1% decline of population mobility was associated with 0.72% (95%CI 0.50% to 0.93%) reduce of cumulative cases for one week, 1.42% two weeks, 1.69% three weeks, 1.72% four weeks,1.64% five weeks and 1.52% six weeks). The impact on weekly new cases seemed greater in the first four weeks, but faded thereafter. The effects on cumulative cases differed by cities of different population sizes, with greater effects seen in larger cities.

**Conclusion:** Persistent population mobility restrictions are well deserved. However, a change in the degree of mobility restriction may be warranted over time, particularly after several weeks of rigorous mobility restriction. Implementation of mobility restrictions in major cities with large population sizes may be even more important.

## Introduction

Since the outbreak of novel coronavirus pneumonia (COVID-19), non-pharmaceutical interventions (NPIs) have been used as a major strategy for mitigating the disease spread.^1,2^ In response to the serious health crisis, China has taken unprecedented measures to contain COVID-19, including Wuhan lockdown and implementation of strict NPIs nationwide.^2^ Those NPIs may generally be classified into two categories. One aimed at controlling sources of infection, including treatment and isolation of confirmed cases, quarantine of exposed persons, or screening of suspicious persons who traveled from Wuhan.^3^ The other was the implementation of human mobility restriction, aiming to address asymptomatic transmission or transmission before symptom onset,^4,5^ and the strategies included suspending intra-city public transport, prohibiting inter-city travels, closing entertainment venues, or banning public gatherings.^2^ Among 342 Chinese cities, 40% suspended intra-city public transport and 64% closed entertainment venues.^3^ The COVID-19 epidemic in China was under control three months after the outbreak.^6,7^

COVID-19 continues to spread across the world.^8^ In the context of resource-limited setting (e.g., lack of virus detection kits), implementing a universal coverage to detect COVID-19 cases is unlikely. The necessary personal protective measures (e.g. face masking) are still in short. As a result, human mobility restriction has been widely used to contain COVID-19 in most countries.^9-11^ However, this strategy has raised extensive criticisms.^12,13^ Emerging studies investigated its effects on containing COVID-19.^3,11,14-16^ including spatial-temporal relation between mobility restriction and diseases transmission^14,15,17^ and initial investigation about the impact of mobility restriction at the early stage. For instance, one study found that suspending intra-city public transport or closing entertainment venues was associated with number of cases reported during the first week of outbreaks;^3^ the other suggested that travel restriction was effective at the early stage of outbreak, but may be less useful when the disease is widespread.^18^ However, distinct knowledge gaps exist about the impacts of human mobility restriction on the disease spread. In particular, the real-world effects of mobility restriction policy remain less clear, as the policy implementation may differ across regions and over time. One plausible argument is that the effects of human mobility restriction may fade over time, and a decision on the level and length of mobility restriction represents a core question for policy makers, whereby disease control and economic growth have to be balanced. All these are linked to casual inferences of the effects of human mobility restrictions and thorough assessments on the magnitude of the effects by time and baseline risks. Thus, we conducted a study to bridge this important gap. The resulting evidence may inform the ongoing global containment of COVID-19.

## Methods

### Design overview

Our study involved two logically linked analyses to quantify the causal effects of human mobility restriction on the spread of COVID-19. Firstly, we applied the difference-in-difference (DID) model to assess real-world effects of mobility restriction policies on mobility declines at the city level. The intra-city population mobility was measured as an index developed by Baidu, a national largest search engine provider in China. Given that Wuhan lockdown was implemented on January 23, we used that date as the starting point for implementing human mobility restriction in China. Subsequently, we used the log-log regression model to examine the effects of mobility declines on the disease spread measured by cumulative or new cases of COVID-19, after adjusting for confounders; the analyses were at the city level given the availability of data and policy implementation. We conducted two analyses to demonstrate the putative causal effects of mobility declines on disease spread measured by number of cumulative cases or weekly new cases.

We collected number of laboratory-diagnosed cases from National Health Commission of China,^7^ Chinese Center for Disease Control and Prevention,^19^ and Provincial Health Commission from January 11 to March 11, 2020, seven weeks after Wuhan lockdown. We chose Jan 11, 2020 as the starting date because the complete genome sequencing of coronavirus was publicly available at that time. This study was approved by the Ethics Review Board of West China Hospital, Sichuan University (2020-99).

### Measure of intra-city population mobility

Intra-city population mobility was measured by a mobility index expressed as an exponentially transformed ratio derived from the daily number of people with outdoor movements divided by the number of residents in a city, and was developed by Baidu, the nationally largest search engine provider in China.^20^ We collected the population mobility index data from Baidu Huiyan system (https://qianxi.baidu.com/). These data were already used for measuring population mobility in previous studies.^18,21-23^

### Effects of mobility restriction policies on population mobility

We used the DID model to examine the causal effects of mobility restriction policies on population mobility at the city level (Supplementary Information Appendix 1). In order to compare the change of population mobility, we paired intra-city mobility index data between 2019 and 2020 by lunar calendar to match the period of the Chinese New Year, during which the world-largest population movements occur. To further adjust for the weekend effect of population mobility, we finally matched Jan 12, 2019 to Jan 4, 2020, both on Saturdays, as the starting date. Through matching, we were able to fairly compare the population movements 24 days before the Chinese New Year and 36 days after between 2019 and 2020. The presence of a parallel trend between 2019 and 2020 confirmed that the assumption of common trends was met for using the DID model.

In the DID model, the effect was expressed as an absolute decline of mobility index at each city between 2020 versus 2019. We also calculated the relative decline of population mobility, expressed as a proportion of the absolute decline by the baseline population mobility index (i. e., that in 2019), and reported relative declines by week (i.e. one, two, three, until six weeks from January 23, 2020) and by population size across cities.^24^

### Impacts of human mobility declines on the spread of COVID-19

We used the log-log regression model to quantify the impacts of human mobility declines on the spread of COVID-19 over time by logarithmic transformation of dependent variables and population declines, controlling for the number of population movements from Wuhan, geo-distances from Wuhan, and population size at each city (Supplementary Information Appendix 1). Using aggregated and anonymized national mobile data (Supplementary Information Appendix 1), we calculated population movements from Wuhan to other cities from Jan 1 to Jan 22, 2020 to resemble the baseline risk (i.e. number of cases imported from Wuhan) in Chinese cities before Wuhan lockdown on Jan 23, 2020.^18^ We counted the number of movements from Wuhan if a person travelled from Wuhan multiple times. Those travelers who stayed in Wuhan for less than two hours were not counted.^25^ We also included other important city characteristics, including population size (in ten thousand unit) and geo-distance from Wuhan (kilometer). Both were collected from Baidu Encyclopedia.

We developed two models to assess the impacts. In the first model, we assessed the effects of accrued mobility declines on cumulative cases, with the exposure of interest expressed as relative declines of population mobility during one week (i.e., Jan 23 to Jan 29), two weeks (i.e., Jan 23 to Feb 5), until six weeks (i.e., Jan 23 to Mar 4). In the second model, we investigated the impact of accrued mobility declines on newly diagnosed cases occurred just one subsequent week (Supplementary Information Appendix 1). Given that the average incubation period of COVID-19 is about 7 days (range 1 to 14 days),^6^ we assumed that the putative effect would occur one week after mobility decline. We additionally explored for potential heterogeneity of effects by population size, measured by number of residents (<1 million, 1 to 5 million, and >5 million).

## Results

### Declines of population mobility among cities in China

Using mobility index data from 344 cities outside Wuhan (4 municipalities, 26 provincial capitals and 314 cities), the average population mobility before January 23, 2020 was comparable to the matched dates in 2019 (Fig.1a). Starting Jan 23, 2020, the population mobility declined dramatically (Fig.1a). The DID model suggested a continual expansion of the relative declines over time in 2020. At the first week, the implementation of mobility restriction policies resulted in 31.35% decline (median, interquartile ranges, −31.35%, −41.63% to −24.27%). After four weeks population mobility declined by 54.81% (−65.50% to −43.56%) (Fig.1a). The relative declines differed by cities, with a larger number of residential populations associated with greater declines (Table 1).

**Fig.1.**
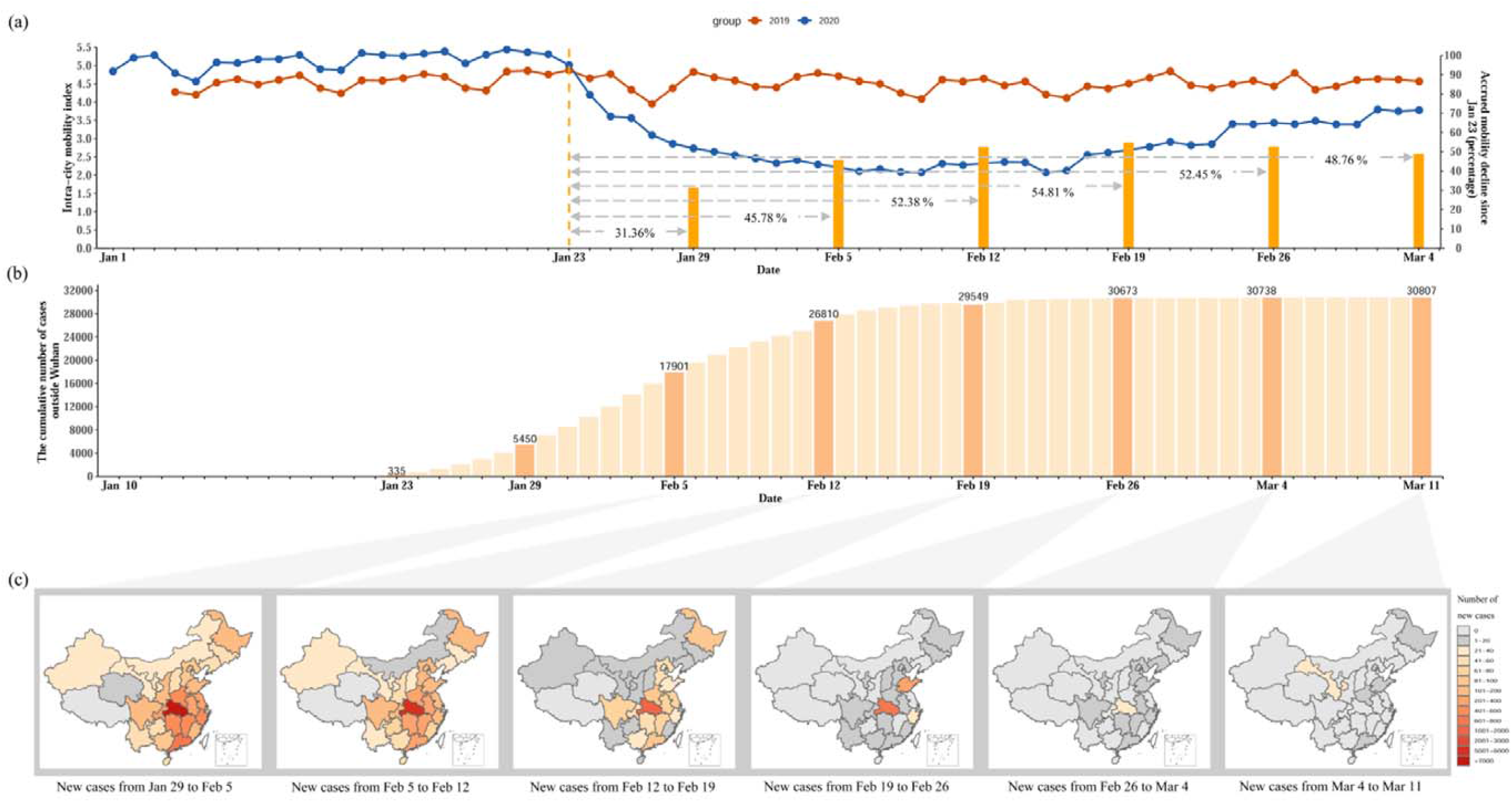
Human mobility declines and the spread of COVID-19 in China. (a) The left vertical axis represents the paralleled trend of average population mobility among cities outside Wuhan between 2019 and 2020 by matched dates, and the right vertical axis indicates the accrued mobility decline since Jan 23. Red dotted line represents the intra-city mobility index in 2020 over dates and the blue dotted line represents the paired intra-city mobility index in 2019 matched by lunar calendar; Yellow dotted line represents the date of Jan 23, 2020. Yellow pillar represents the relative decline in population mobility between 2020 and 2019 over weeks, calculated by DID. Horizontal axis indicates the dates from Jan 1 to Mar 4, 2020. (b) The cumulative number of cases diagnosed in cities outside Wuhan over dates. (c) The number of new cases diagnosed in provinces (excluded Wuhan in Hebei province) over weeks.

**Table 1.** Relative declines in population mobility between 2020 and 2019 (percentage)

| Duration of policy implementation | Number of cities | Mean (Standard deviation) | Median (Percentile 25- 75) |
| --- | --- | --- | --- |
| One week (Jan 23 - Jan 29, 2020) |  |  |  |
| Number of residents | 344 | -33.54 (13.88) | -31.35 (-41.63 - -24.27) |
| <1 million | 42 | -22.27 (13.70) | -20.57 (-28.39 - -12.18) |
| ≥1 million and < 3 million | 112 | -31.03 (11.48) | -29.89 (-36.30 - -22.94) |
| ≥3 million and <5 million | 92 | -33.39 (10.71) | -32.37 (-39.62 - -22.50) |
| ≥5 million and <10 million | 83 | -39.27 (13.78) | -39.49 (-47.24 - -28.48) |
| ≥10 million | 15 | -52.98 (15.30) | -48.89 (-61.46 - -42.60) |
| Two weeks (Jan 23 - Feb 5, 2020) | 344 | -47.59 (14.75) | -45.78 (-56.10 - -37.19) |
| <1 million | 42 | -38.38(15.32) | -36.93 (-49.03 - -25.44) |
| ≥1 million and < 3 million | 112 | -44.98 (11.53) | -42.58 (-51.07 - -36.81) |
| ≥3 million and <5 million | 92 | -46.81 (11.85) | -45.56 (-54.59 - -38.08) |
| ≥5 million and <10 million | 83 | -52.81 (16.06) | -53.02 (-62.10 - -41.17) |
| ≥10 million | 15 | -68.76 (14.36) | -65.20 (-80.15 - -60.10) |
| Three weeks (Jan 23 - Feb 12, 2020) | 344 | -53.10 (15.28) | -52.38 (-62.51 - -42.10) |
| <1 million | 42 | -46.11(16.75) | -46.25 (-57.10 - -35.03) |
| ≥1 million and < 3 million | 112 | -50.52 (11.92) | -48.16 (-57.40 - -42.01) |
| ≥3 million and <5 million | 92 | -52.00 (12.85) | -51.37 (-60.40-41.99) |
| ≥5 million and <10 million | 83 | -57.58 (17.21) | -56.82 (-67.50 - -46.77) |
| ≥10 million | 15 | -73.93 (13.16 ) | -71.72 (-82.98 - -66.02) |
| Four weeks (Jan 23 - Feb 19, 2020) | 344 | -54.65 (15.60) | -54.81 (-65.50 - -43.56) |
| <1 million | 42 | -50.05 (18.45) | -51.24 (-62.01 - -38.43) |
| ≥1 million and < 3 million | 112 | -52.07 (12.45) | -49.77 (-61.64 - -44.04) |
| ≥3 million and <5 million | 92 | -53.43 (13.62) | -52.94 (-73.03 - -42.99) |
| ≥5 million and <10 million | 83 | -58.14 (17.25) | -57.95 (-69.71 - -46.60) |
| ≥10 million | 15 | -75.01 (12.18) | -73.87 (-82.78 - -66.97) |
| Five weeks (Jan 23 - Feb 26, 2020) | 344 | -52.48 (16.20) | -52.45 (-64.70 - -40.72) |
| <1 million | 42 | -51.21 (20.10) | -51.19 (-67.38 - -36.28) |
| ≥1 million and < 3 million | 112 | -49.80 (13.61) | -48.56 (-60.20 - -39.90) |
| ≥3 million and <5 million | 92 | -50.82 (14.51) | -50.35 (-58.05 - -40.03) |
| ≥5 million and <10 million | 83 | -54.89 (17.15) | -57.66 (-66.44 - -42.72) |
| ≥10 million | 15 | -72.85 (11.57) | -73.52 (-81.29 - -63.88) |
| Six weeks (Jan 23 - March 4, 2020) | 344 | -49.44 (16.67) | -48.76 (-61.18 - -36.91) |
| <1 million | 42 | -50.52 (21.38) | -49.54 (68.51 - -32.81) |
| ≥1 million and < 3 million | 112 | -46.81 (14.68) | -44.38 (-55.85 - -36.25) |
| ≥3 million and <5 million | 92 | -47.44 (15.03) | -46.82 (-55.23 - -36.37) |
| ≥5 million and <10 million | 83 | -51.11 (16.82) | -53.67 (-62.91 - -38.12) |
| ≥10 million | 15 | -69.13 (10.91) | -70.05 (-79.26 - -59.61) |

### Effects of human mobility declines on the spread of COVID-19

There were 335 cases diagnosed in cities outside Wuhan by Jan 23, at Wuhan lockdown. Four weeks later, the cumulative number of cases stabilized, and then a small number of cases increased thereafter. By Mar 11, a total of 30807 COVID-19 cases were diagnosed in cities outside Wuhan of Mainland China (Fig.1b). The new cases of COVID-19 were rapidly reduced over weeks (Fig.1c). We also found the potential linear trend between the relative declines of population mobility and numbers of cumulative and new COVID-19 cases over time by population size of cities (Fig.2). Table S1 reported the median population size in cities (median 3 360 000, interquartile ranges, 1 837 700 to 5 320 400), geo-distance from Wuhan (844.00 kilometers, 561.50 - 1291.50), and number of movements from Wuhan before Wuhan lockdown (2858 times, 661 - 9305).

**Fig.2.**
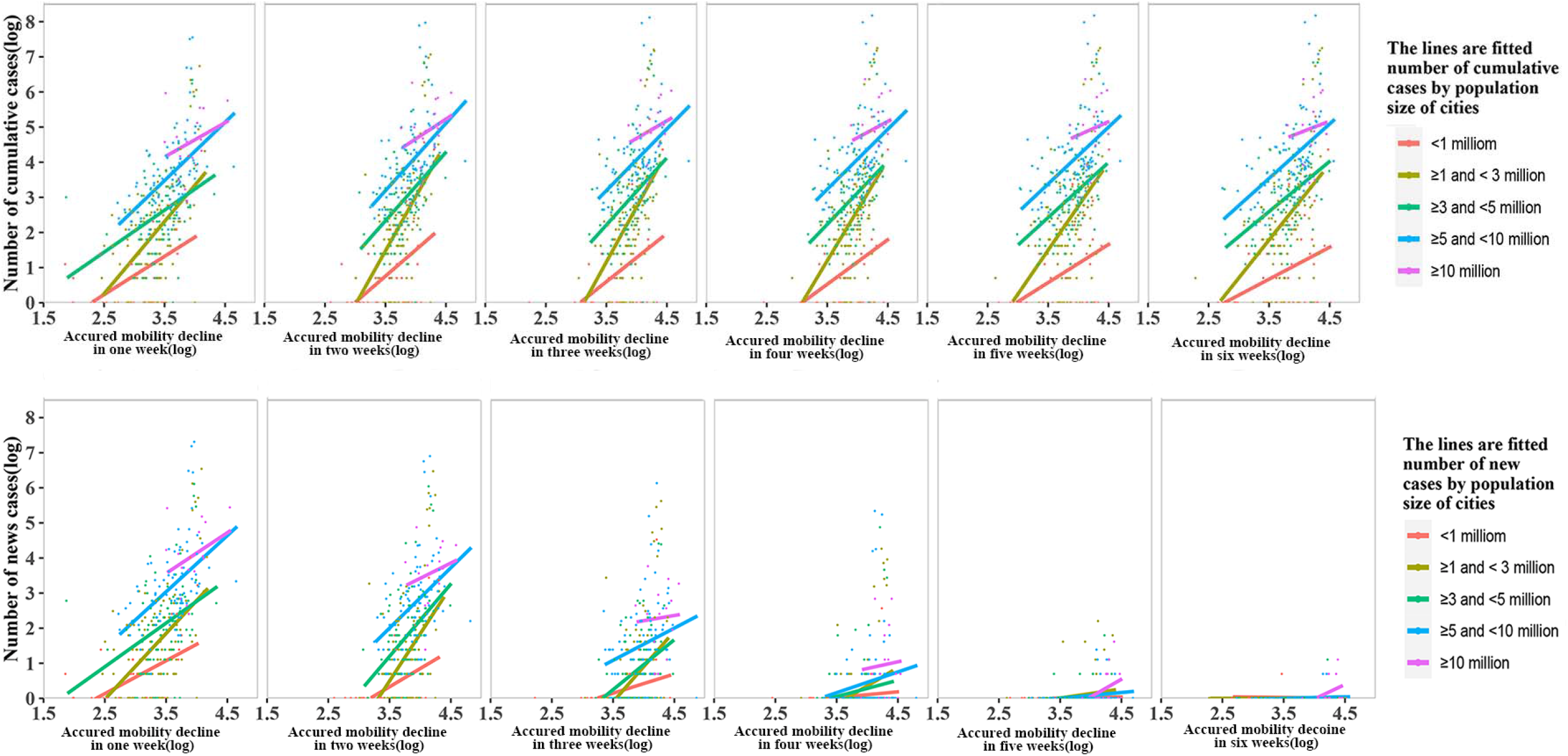
Linear trend between the relative declines of population mobility (logarithmic transformation) and numbers of cumulative and new COVID-19 cases (logarithmic transformation) over weeks by population size of cities.

The log-log regression models reported the effects of accrued mobility declines on cumulative cases (Fig.3). At one week, a 1% decline of population mobility was associated with 0.72% (95%CI 0.50% to 0.93%) reduction in number of cumulative cases, after adjusting for other confounders. The magnitude of effects continued to increase until four weeks (1.42%, 1.11% to 1.74% for two weeks; 1.69%, 1.34% to 2.03% three weeks; 1.72%, 1.38% to 2.05% four weeks; 1.64%, 1.35% to 1.94% five weeks and 1.52%, 1.25% to 1.78% six weeks). However, the effects of mobility declines differed among cities of varying population sizes, with larger effects among larger cities, as opposed to smaller ones (less than 1 million) (Fig.4).

**Fig.3.**
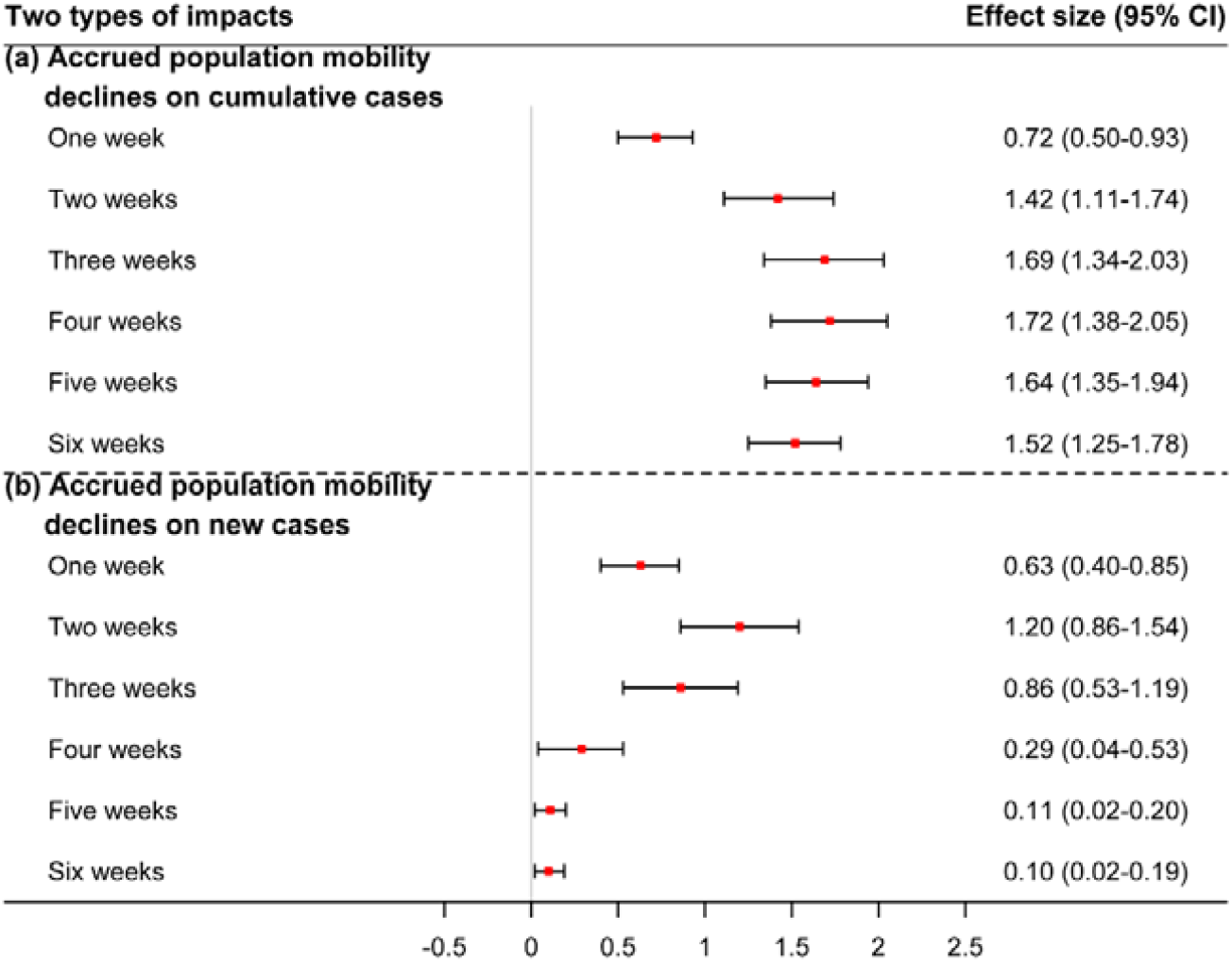
The effects of accrued mobility declines on cumulative cases. Effect size represents the elasticity (i.e., %Δy/%Δx) of the number of COVID-19 cases to population mobility decline.

**Fig.4.**
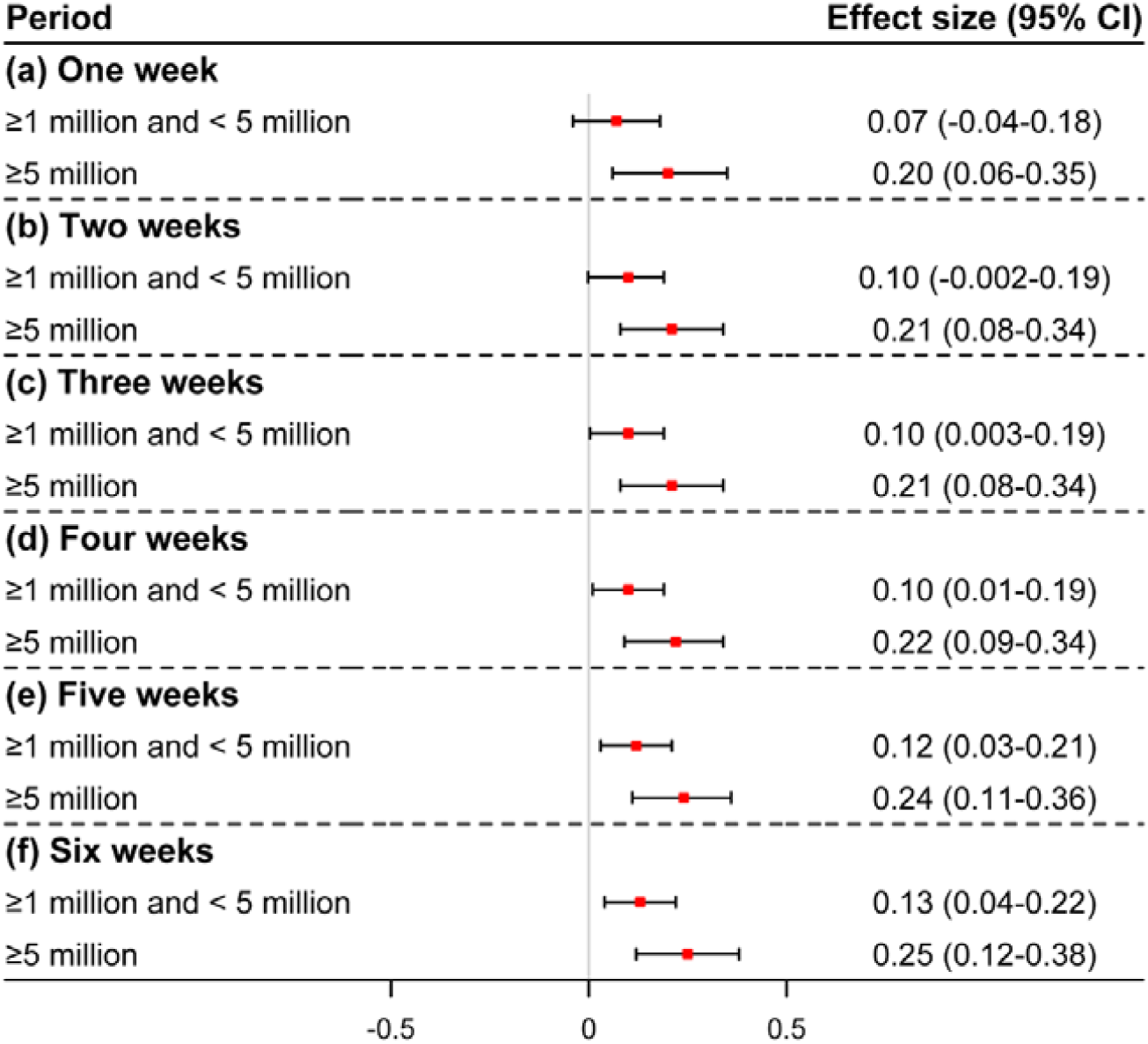
Heterogeneous effects of accrued mobility declines on cumulative cases by city scale. Effect size represents the elasticity (i.e., %Δy/%Δx) of the number of COVID-19 cases to population mobility decline.

There also were statistically significant associations between accrued mobility declines and new cases occurred at the subsequent week. At two weeks, a 1% decline in population mobility corresponded to 1.20% (95%CI 0.86% to 1.54%) reduction in new cases at the subsequent week (i.e., third week). Even after six weeks, the effect was observed, although small (Fig.3). No heterogeneity of effects was observed in this model (Table S2).

## Discussion

### Summary of our findings

Human mobility restriction has ever been used at the outbreak of fulminating infectious diseases, such as SARS in 2003^26,27^ and influenza A in 2009,^28,29^ but with a narrower scope or a transitory period.^28,30,31^ The outbreak of COVID-19 in China, which coincided with the world largest population movements surrounding Chinese New Year (i.e., *Chun Yun*), offered an unprecedented opportunity to investigate the real-world effects of population mobility restrictions.

By the counter-fact DID model, we found almost one third of mobility declines occurred in the first week after the policy implementation; after four weeks, the population mobility lowered by 54.8%. Policies’ impacts on mobility decline varied by population size of a city and the effects persisted over time. In our critical analyses that assessed the effects of mobility declines on the disease spread, we found that the accrued population declines were associated with the reduction of both cumulative CVID-19 cases and new cases across the six weeks after the policy implementation. More interestingly, we found that its impact on new cases seemed larger in the first four weeks, and faded after five weeks.

So, what do these findings tell us? First of all, we believe that the implementation of population mobility restrictions is appropriate and strongly needed, in balancing the serious pandemic and the fatal consequences it has caused versus the economic loss and temporary loss of personal mobility. Up to March 11, 2020, a total of 30 documents were issued from national governmental authority (Table S3). These policy documents covered nearly all social activities across cities and rural areas, including healthcare services, education, transportation, tourism, elderly care, child welfare service, employment, work at home, and community management. Accordingly, the continual policies implementation resulted in the promptly mobility declines, and further lead to effective and rapid reduction of new cases in subsequent weeks, which would, in turn, rapidly eliminate the impact of the pandemic. Secondly, we may also infer from our findings that the effects of the mobility restriction policies would be large in the first few weeks, but attenuate over time. After five weeks, their effects became smaller. This alerts that persistent mobility restrictions are highly deserved, but flexible mobility restriction policies may be warranted particularly after several weeks of rigorous mobility restriction. Thirdly, our study suggested larger effects among bigger cities over time, which highlighted that the implementation of mobility restrictions in major metropolises is particularly important and meaningful. Fourthly, the findings strongly advised that, in resource-limited setting where healthcare resources are not readily available, non-medical interventions may be optimal strategies in containing the disease transmission. We also believe that these strategies are not just effective in the early stage of the disease outbreak, but also at the stage of wide spread, because of its sheer mechanism to block population contacts at emergencies. Thus, those nations amid the disease pandemic may still consider implementing these strategies.

### Comparison with previous studies

As an initial mobility restriction intervention, Wuhan lockdown was extensively assessed.^3,32-35^ Previous studies showed that the Wuhan lockdown delayed the arrival of COVID-19 in other Chinese cities by 2.9 days,^3^ and lowered 64.8% of cases in 347 Chinese cities outside Hubei.^33^ However, a large number of movements actually occurred before Wuhan lockdown, and such populations were likely infected or even symptomatic that drove the number of cases identified in cities outside Wuhan.^3^

Several studies investigated the effects of other measures,^3,11,14-16,36-38^ among which simulations were extensively used to quantify the effects of physical distancing, early case detection or isolation, or combination of multiple measures.^36-38^ These studies provided important insights about the impacts of mobility restriction measures, but had limitation given the use of modelling, whereby assumptions are often employed.

Up to now, empirical studies still fall short.^39^ One early study assessed the impacts of human mobility restriction on COVID-19 cases at the first week, and the other suggested that travel restriction was more useful in the early outbreak, but attenuated if the outbreak was expanded. However, the extent to which the mobility restriction policy led to mobility decline and whether human mobility restriction causally controlled the spread of COVID-19 were not yet established. An earlier study, using mobility data from four metropolitan areas in USA, mainly examined the temporal correlation between timing of public policy measures and cumulative cases of COVID-19,^17^ but indicated a lack of causality due to the nature of descriptive analyses.^17^

### Strengths and limitations

Our study has several strengths. Firstly, we have used rigorous methods to assess the causal effects of human mobility restriction on the spread of COVID-19. We have also profiled the declines of population mobility by using the DID model, which avoided the reverse causality and confounding by usual fluctuation of population mobility over time. Secondly, we included important confounders in the models with precise measurements, such as number of times of population movements from Wuhan to imported regions, number of residents (ten thousand) and geo-distance from Wuhan (kilometer). As a result, good model fitting is achieved (*R*^2^ > 70%). Thirdly, we have used thorough and real-time population mobility data to assess the impacts of non-pharmaceutical interventions during the outbreak of infectious diseases. Real-time mobility data, such as airline flights data^40,41^ and aggregated human mobility data,^25,42^ when combining routine epidemiological surveillance, could play a crucial role during the disease pandemic.

Our study had limitations. Firstly, the measurement of population mobility may not be optimal due to the fact that not everyone uses Baidu App in their smartphones and some subgroup population (e.g. children or elderly) were not covered. Nevertheless, it covers about 55% of smartphones users in China,^23^ and was well used previously. Thus, we believe that it is nationally representative likely exist. Secondly, in our analyses, we assumed that the mobility restriction would be effective one week after given the reported incubation period between 1 to 14 days.^6^ This assumption may affect the resulting estimates, and slight changes may occur.

## Conclusion

In response to COVID-19 epidemic, China has implemented a comprehensive set of mobility restriction policies, which resulted in more than 50% population mobility declines just in a few weeks; the effect was more pronounced in large cities and remained consistent over time. The resulting population mobility declines had a direct impact on the reduction of cumulative and new COVID-19 cases; this effect particularly had larger effect in the first few weeks and attenuated thereafter, and was also more pronounced in larger cities. Our study confirmed that strict implementation of comprehensive population mobility restriction policies was highly warranted, particularly in first several weeks and the large cities.

## Data Availability

The data about population mobility, daily and cumulative COVID-19 cases at city level in mainland China, and outbound flow data from Wuhan to all other municipal cities during January 1, 2020 to January 22, 2020, may be available with the approval by the research team.

## Funding

This work was supported by National Natural Science Foundation of China (71704122,71974138), and the National Science and Technology Major Project (2018ZX10302206), 1·3·5 project for disciplines of excellence, West China Hospital, Sichuan University (ZYYC08003).

## Acknowledgements

None.

## Author Contributions

Jing Tan, Xin Sun and Weimin Li conceptualized the study. Jing Tan analyzed and interpreted data, wrote the first draft of the article, and contributed to all revisions. Yiquan Xiong collected the data and co-wrote the first draft. Shaoyang Zhao analyzed and interpreted data and revised the draft critically for important intellectual content. Chunrong Liu and Shiyao Huang collected the data. Xin Lu interpreted data. Lehana Thabane and Feng Xie revised the draft critically for important intellectual content.

## Competing of interests

None declared

## Supplementary Information

## Appendix 1

### Methods

#### Characteristics of imported cities

The population movements data was aggregated and anonymized at the county-level from China Unicom, one of largest mobile operators in China. In the efforts to enhance the extrapolation of data, the operator helped us use a recognized machine learning algorithm, involving the users’ age, gender, operator’s coverage and other parameters, to arrive at the whole network coverage with all users. The same data resource has been used in previous study [1].

[1] Jia JS, Lu X, Yuan Y, Xu G, Jia J, Christakis NA. Population flow drives spatio-temporal distribution of COVID-19 in China. Nature. 2020.

#### The effects of mobility restriction policies on population mobility

The DID model could be described as follows.

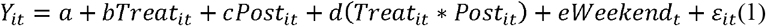

Where *i, t* respectively index the city and date, and *Y*_*it*_ is the dependent variable representing the population mobility in the *i* city at *t* date. ***T****reat*_*it*_ is a dummy variable, setting 0 for the status of 2019, where no human mobility restriction implemented, as the control group; 1 for the status in 2020, where human mobility restriction had been implemented in response to outbreak, as the intervention group. ***P****ost*_*it*_ is a dummy, taking 0 for the period before lockdown of Wuhan (e.g. before January 23 in 2020 or before February 3 in 2019) and 1 for the period after that. ***W****eekend*_***t***_ is a dummy measuring whether the *t* date is a weekend. The coefficient d of interaction term, multiplying ***T****reat*_*it*_ by ***P****ost*_*it*_, is the effect value to be estimated, which is the absolute decline of population activity in *i* city at *t* date of 2020, compared with those in 2019. We further calculated a relative change of population mobility in 2020 over dates, with the absolute decline of population mobility in 2020 divided by average population mobility at the corresponding period in 2019.

#### Impacts of human mobility restriction on the spread of COVID-19

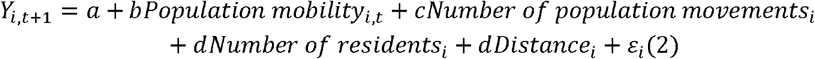

Where *i* index the city, *t* index week (*t = 1,2,3,4,5,6*), and *Y*_*i,t+1*_ is the dependent variable representing the number of diagnosed cases in *i* city at time *t+1*. ***P****opulation mobility*_*i*_ is on behalf of the relative declines of population mobility in *i* city during time *t, Number of population movements*_*i*_ means the number of times of population movements from Wuhan to imported regions before January 23, 2020 in *i* city. *Number of residents*_*i*_ is the number of residents in *i* city, *Distance*_*i*_ is the geo-distance from Wuhan to *i* city. The dependent variable of *Y*_*i, t +*1_ and population activities were undertaken logarithmic transformation. Given that the average incubation period of COVID-19 is about 7 days (range 1 to 14 days), we assumed that the putative effect would occur one week after mobility decline. The coefficient of *b* thus represents the elasticity (i.e., %*Δ*y/%*Δ*x) of the number of COVID-19 cases to population mobility decline.

**Table S1.** The characteristics of included cities in models*. *We totally included 344 cities in China

| Variable | Mean (Standard deviation) | Median (Percentile 25-Percentile 75) |
| --- | --- | --- |
| Change of population mobility (%) |  |  |
| One week (Jan 23 - Jan 29, 2020) | -33.54 (13.88) | -31.35 (-41.63 - -24.27) |
| Two weeks (Jan 23 - Feb 5, 2020) | -47.59 (14.75) | -45.78 (-56.10 - -37.19) |
| Three weeks (Jan 23 - Feb 12, 2020) | -53.10 (15.28) | -52.38 (-62.51 - -42.10) |
| Four weeks (Jan 23 - Feb 19, 2020) | -54.65 (15.60) | -54.81 (-65.50 - -43.56) |
| Five weeks (Jan 23 - Feb 26, 2020) | -52.48 (16.20) | -52.45 (-64.70 - -40.73) |
| Six weeks (Jan 23 - March 4, 2020) | -49.44 (16.67) | -48.76 (-61.18 - -36.91) |
| Number of residents (thousand) | 4099.10 (3481.20) | 3360.00 (1837.70 - 5320.40) |
| Number of population movements (time) | 55553 (3385367) | 2858 (661 - 9305) |
| Distance from Wuhan (kilometer) | 1033.97 (701.52) | 844.0 (561.50 - 1291.50) |

\*We totally included 344 cities in China

**Table S2.** The heterogeneity of impacts of accrued mobility declines and new cases occurred at the subsequent week by city scale (coefficients for interaction terms, 95%CI)

| Period | Number of cases | Effect size |
| --- | --- | --- |
| One week (Jan 23 - Jan 29, 2020) - new cases from Jan 29 to Feb 5 |  |  |
| ≥1 million and < 5 million |  | 0.01 (-0.11-0.13) |
| ≥5 million |  | 0.16 (0.009-0.32) |
| Two weeks (Jan 23 - Feb 5, 2020) - new cases from Feb 5 to Feb 12 |  |  |
| ≥1 million and < 5 million |  | 0.01 (-0.09-0.12) |
| ≥5 million |  | 0.11 (-0.03-0.25) |
| Three weeks (Jan 23 - Feb 12, 2020) - new cases from Feb 12 to Feb 19 |  |  |
| ≥1 million and < 5 million |  | -0.04 (-0.13-0.05) |
| ≥5 million |  | 0.0004 (-0.12-0.13) |
| Four weeks (Jan 23 - Feb 19, 2020) - new cases from Feb 19 to Feb 26 |  |  |
| ≥1 million and < 5 million |  | -0.07 (-0.13-0.002) |
| ≥5 million |  | -0.14 (-0.24- -0.05) |
| Five weeks (Jan 23 - Feb 26, 2020) - new cases from Feb 26 to Mar 4 |  |  |
| ≥1 million and < 5 million |  | -0.008 (-0. 04-0.02) |
| ≥5 million |  | -0.05 (-0.09- -0.007) |
| Six weeks (Jan 23 - March 4, 2020) - new cases from Mar 4 to Mar 11 |  |  |
| ≥1 million and < 5 million |  | -0.006 (-0.02-0.005) |
| ≥5 million |  | -0.01 (-0.03-0.005) |

\*Cities with number of residents less than 1 million were deemed as reference.

**Table S3.** Human mobility restriction documents issued from national governmental authorities.

| Order | Issued time | Government organization | Title | Main contents |
| --- | --- | --- | --- | --- |
| 1 | 20-Jan | National Health Commission of the People's Republic of China | National Health Commission of the People's Republic of China Held a National Teleconference to Deploy the Prevention and Control Strategy of COVID-19 | Strengthening the population activity management in Hubei province and Wuhan city, and reducing the large-scale public gathering |
| 2 | 20-Jan | National Health Commission of the People's Republic of China | COVID-19 Has Been Included in Notifiable Disease Management | COVID-19 has been classified as Class B infectious disease and managed as Class A infectious disease |
| 3 | 21-Jan | Ministry of Education of the People's Republic of China | Notice of Carefully Taking Precautionary Measures to Prevent and Control COVID-19 in the Education System | 1. The Ministry of Education requires that educational units at all levels cannot organize aggregated activities if without permit;<br>2. Ensuring educational units at all levels have implemented prevention and control measures of COVID-19 in schools effectively. |
| 4 | 21-Jan | National Railway Administration of People's Republic of China | National Railway Administration of People's Republic of China Has Implemented the Prevention and Control Measures to Ensure Smooth and Orderly Spring Festival Transportation | From January 24th 0:00 on, the free railway ticket refund fee privilege will be extended to cover all passengers who bought railway tickets (passengers who planned to go to Hubei province only before), aiming to encourage people to refund and to reduce travel. |
| 5 | 22-Jan | The General Office of Ministry of Culture and Tourism of the People's Republic of China;<br>The General Office of National Cultural Heritage Administration | Notice of Carefully Taking Precautionary Measures to Prevent and Control COVID-19 | Public cultural service units at various places are required to limit the number and scale of mass cultural activities strictly and reduce the public gathering. |
| 6 | 23-Jan | Civil Aviation Administration of China | Notice on Waiver of Air Ticket Refund Fee | From January 24th 0:00 on, the free air ticket refund fee privilege will be extended to cover all passengers who bought air tickets (passengers who planned to go to Hubei province only before), aiming to encourage people to refund and to reduce travel. |
| 7 | 23-Jan | Ministry of Education of the People's Republic of China | Ministry of Education deploys the prevention and control emergency plans of COVID-19 on education system | 1. The administrative departments and schools at all levels of education do not, in principle, held large-scale, aggregated activities and examinations;<br>2. Schools at all levels should postpone original enrollment plans and examinations;<br>3. The primary and secondary school students are not allowed to participate in competitions, talent shows and other large-scale gathering activities and examinations organized by social institutions;<br>4. Universities in Wuhan and other areas with high COVID-19 epidemic should strictly control outsiders to enter campus at will, and cancel all kinds of tourist groups to enter the campus. |
| 8 | 24-Jan | The General Office of Ministry of Culture and Tourism of the People's Republic of China | Urgent Notice of Suspending the Business Activities of Tourism Enterprises to Prevent and Control COVID-19 | From January 24th on, travel agencies and online travel companies suspended providing group travel services and "ticket & hotels" tourism products nationwide. |
| 9 | 24-Jan | A Central Leadership Group for Epidemic Response and the Joint Prevention and Control Mechanism of the State Council | Notice on Strengthening the Prevention and Control of COVID-19 in Communities | According to the cases number, community was classified as no cases community, sporadic cases community and outbreak community. The former two types of community should strengthen health education to guide residents to reduce travel and avoid aggregation activities, while the outbreak community should implement closed management strategy to limit people aggregation. |
| 10 | 26-Jan | National Health Commission of the People's Republic of China | Notice on Strengthening the Prevention and Control of COVID-19 in Primary Medical and Health Institutions | 1. Training the primary medical staffs about the COVID-19 by the means of Internet;<br>2. Contacting with the management objects by the means of family doctor APP, cable television network, WeChat and so on. |
| 11 | 26-Jan | The General Office of the State Council of the People's Republic of China | Notice of Extending the Spring Festival Vacation in 2020 | 1. Extending the 2020 Spring Festival vacation to February 2nd (originally scheduled for January 31th).<br>2. Postponing the 2020 Spring Semester Starting Times, which specific time is notified by the Ministry of Education. |
| 12 | 27-Jan | Ministry of Education of the People's Republic of China | Notice of Postponing the Spring Semester Starting Times in 2020 | 1. Postponing the 2020 spring semester starting times and students are not allowed to return schools in advance without approval;<br>2. Students are required to keep staying at home, and stop holding and participating in party or any other gathering activities during winter holidays. |
| 13 | 28-Jan | Ministry of Civil Affairs of the People's Republic of China | Urgent Notice of Requiring All Localities to Do Their Best to Prevent and Control COVID-19 in Child Welfare Service Institutions | 1. Implementing Closed Management Strategy in child welfare services institutions.<br>2. Canceling all outdoor activities and indoor gathering activities. |
| 14 | 28-Jan | A Central Leadership Group for Epidemic Response and the Joint Prevention and Control Mechanism of the State Council | Notice of Carefully Taking Precautionary Measures to Prevent and Control COVID-19 for the Elderly People | 1. Service agencies for the elderly should avoid the gathering and collective activities of the elderly as much as possible;<br>2. Implementing Closed Management Strategy in pension agencies when necessary;<br>3. Strengthening the health education for the rural left-behind elderly carefully. |
| 15 | 28-Jan | National Health Commission of the People's Republic of China | Notice of Carefully Taking Precautionary Measures to Prevent and Control COVID-19 in Childcare Institution | 1. All kinds of foster care institutions can suspend caring services in accordance with the related laws;<br>2. The early-stage education institutions and parent-child gardens that provide services for children aged 3 and below should stop live training activities. |
| 16 | 28-Jan | A Central Leadership Group for Epidemic Response and the Joint Prevention and Control Mechanism of the State Council | Notice of Issuing the Recent Prevention and Control COVID-19 Working Scheme | 1. Minimizing the movement of people, encouraging the public to keep themselves staying at home and reducing visits to friends and relatives;<br>2. Further strengthening the management of large-scale public gathering activities, canceling or postponing all kinds of large-scale activities, and closing cultural tourism facilities;<br>3. Limitation on gathering meals during the Chinese New Year vacation. |
| 17 | 29-Jan | Ministry of Civil Affairs of the People's Republic of China | The Ministry of Civil Affairs Has Required the Civil Affairs System at All Levels to Thoroughly Study the Spirit of the Series of Crucial Instructions about the COVID-19 of General Secretary Xi Jinping and has encouraged them to do all that they can to Prevent and Control COVID-19 | 1. Strengthening the closed management to foster care institutions, and the entry and exit of unnecessary persons shall be strictly prohibited;<br>2. Reducing staff-intensive meetings and canceling unnecessary research activities. |
| 18 | 29-Jan | Ministry of Civil Affairs of the People's Republic of China | Notice of Issuing the Implementation Scheme for COVID-19 Prevention and Control of Social Organization Registration Management Institution | 1. To cancel or postpone, as far as possible, meetings and activities that are has no relationship with the COVID-19 epidemic, such as training and external exchanges;<br>2. To motivate people to consult and work online or by means of telephone, so as to minimize face-to-face contact. |
| 19 | 29-Jan | Ministry of Civil Affairs of the People's Republic of China;<br>National Health Commission of the People's Republic of China | Urgent Notice of Further Motivating Urban and Rural Community Members Carrying Out COVID-19 Prevention and Control Working | 1. No gathering activities are allowed in communities before the end of the COVID-19 epidemic;<br>2. Specific places in community are not allowed to provide gathering services to the public, such as laces that providing gathering services such as community library, entertainment room, senior activity center and so on;<br>3. Persuading community groups to suspend business when necessary |
| 20 | 29-Jan | The General Office of National Health Commission of the People's Republic of China;<br>The General Office of National Administration of Traditional Chinese Medicine | Notice of Carefully Coping With the Healthcare Utilization Peak Situation After the 2020 Spring Festival Vacation | 1. Encouraging conditional medical institutions (i.e. medical institutions that has enough medical staffs) to provide online or telephone consultation services, or appropriately extend the prescription dosage for patients with chronic and geriatric diseases to reduce the number of visits to hospitals;<br>2. Guidance for patients on use of healthcare services at different time, dissuading people from going to hospitals if the situation is not serious, and reducing patient's aggregation as far as possible. |
| 21 | 30-Jan | A Central Leadership Group for Epidemic Response and the Joint Prevention and Control Mechanism of the State Council | Notice on Calling for More Efforts to Strengthen Prevention and Control of COVID-19 in Rural Areas | 1. Suspending any ethnographical activities for Spring festival, such as garden fate and temple fair;<br>2. Persuading the rural residents not to held large-scale activities, such as wedding and funeral;<br>3. Encouraging the rural residents to stay at home, avoiding to visit relatives and friends. |
| 22 | 31-Jan | Ministry of Veterans Affairs of the People's Republic of China | Notice on the Tomb-sweeping for the Martyrs in the Epidemic Control of COVID-19 | 1 All of Martyrs institutions should be temporarily closed, the opening time will be decided and noticed depending on epidemic situation;<br>2. All of Martyrs institutions and must not organize any clustering commemorative activities. |
| 23 | 2-Feb | National Health Commission of the People's Republic of China | Notice on Urging Prevention and Control of COVID-19 Outbreak among Children and Pregnant Women | Suspending or avoiding some unnecessary face-to-face diagnosis and treatment services for children and pregnant women, and strengthening the online medical consultation and guidance. |
| 24 | 3-Feb | National Health Commission of the People's Republic of China | Notice on Strengthening Informatization to Support the Prevention and Control of COVID-19 Outbreak | 1. Actively organizing medical institutions at all levels to use the "Internet +" to provide medical services, such as online voluntary consultation about COVID-19, and guidance for medical observation at home;<br>2. Taking advantages of Internet hospitals and Internet diagnosis and treatment services, and encouraging the online diagnosis of some common and chronic diseases and drug delivery services;<br>3. Strengthening the Internet government services. |
| 25 | 4-Feb | A Central Leadership Group for Epidemic Response and the Joint Prevention and Control Mechanism of the Ministry of Education | Guidance on the Organization and Management of online teaching among Ordinary Colleges and Universities | Suspending all winter vacation social practice in colleges and universities, and the planned winter vacation social practice should be carried over to the next year or next semester. |
| 26 | 6-Feb | The General Office of Ministry of Human Resources and Social Security of the People's Republic of China | Notice on Management of Human Resources Market in the Epidemic Control of COVID-19 | 1. Suspending on-site employment activities;<br>2. Strengthening online employment and other online activities. |
| 27 | 6-Feb | The General Office of Ministry of Commerce of the People's Republic of China;<br>The General Office of National Health Commission of the People's Republic of China | Notification on Promoting Prevention and Control of COVID-19 Outbreak in Living Service Enterprises | 1. Don't organize large-scale product promotion, exhibition and sales activities;<br>2. It is strictly forbidden to open business establishments with the high risk of transmission of COVID-19 |
| 28 | 6-Feb | The General Office of National Health Commission of the People's Republic of China | Notification on Promoting Online Consulting Services for Diagnosis and Treatment by Internet in the Epidemic Control of COVID-19 | 1. Provincial health administrative departments should establish unified provincial internet medical service platforms and service management platform for prevention and control of the COVID-19;<br>2. Strengthening medical institutions with the online platforms to conduct online service for COVID-19, and guiding patients to seek medical treatment in an orderly manner. |
| 29 | 7-Feb | Ministry of Human Resources and Social Security of the People's Republic of China;<br>China Enterprise Confederation;<br>China Enterprise Directors Association;<br>All-China Federation of Industry and Commerce | Opinions on Strengthening of Stabilizing Labor Relations and Supporting Resumption of Enterprises in the Epidemic Control of COVID-19 | 1. Encouraging enterprises to make arrangements for employees to work at home, except for employees of enterprises related to disease control and prevention;<br>2. Encouraging resumption enterprises to implement flexible employment measures, such as commuting at different times and flexible commuting. |
| 30 | 24-Feb | A Central Leadership Group for Epidemic Response and the Joint Prevention and Control Mechanism of the State Council | Notice on Urging Prevention and Control of COVID-19 outbreak in Scientifically and Accurate Way | 1. The campus implements closed management, and students are strictly forbidden to return to school in advance without permission;<br>2. Strengthening the prevention and control of COVID-19 in rural area, and reducing the crowd gathering activities (e.g. fairs) |

## Notes

### Competing Interest Statement

The authors have declared no competing interest.

### Clinical Trial

This is not a clinical trial.

### Author Declarations

This study was approved by the Ethics Review Board of West China Hospital, Sichuan University (2020-99)

## Reference

1. Anderson RM, Heesterbeek H, Klinkenberg D, Hollingsworth TD. How will country-based mitigation measures influence the course of the COVID-19 epidemic? Lancet. 2020;395:931–4.

2. Cowling BJ, Aiello A. Public health measures to slow community spread of COVID-19. J Infect Dis. 2020;221:1749–51.

3. Tian H, Liu Y, Li Y, et al. An investigation of transmission control measures during the first 50 days of the COVID-19 epidemic in China. Science. 2020;368:638–42.

4. Nishiura H, Kobayashi T, Suzuki A, et al. Estimation of the asymptomatic ratio of novel coronavirus infections (COVID-19). Int J Infect Dis. 2020;94:154–5.

5. Hellewell J, Abbott S, Gimma A, et al. Feasibility of controlling COVID-19 outbreaks by isolation of cases and contacts. Lancet Glob Health. 2020;8:e488–e96.

6. WHO, Report of the WHO-China Joint Mission on Coronavirus Disease 2019 (COVID-19). 2020. https://www.who.int/docs/default-source/coronaviruse/who-china-joint-mission-on-covid-19-final-report.pdf (28 April 2020, date last accessed).

7. National Health Commission of the People’s Republic of China. http://www.nhc.gov.cn/xcs/yqtb/list_gzbd.shtml (28 April 2020, date last accessed).

8. WHO, Coronavirus disease 2019 (COVID-19) Situation Report – 73. 2020. https://www.who.int/docs/default-source/coronaviruse/situation-reports/20200402-sitrep-73-covid-19.pdf?sfvrsn=5ae25bc7_2 (28 April 2020, date last accessed).

9. The L. India under COVID-19 lockdown. Lancet. 2020;395:1315.

10. Tobias A. Evaluation of the lockdowns for the SARS-CoV-2 epidemic in Italy and Spain after one month follow up. Sci Total Environ. 2020;725:138539.

11. Signorelli C, Scognamiglio T, Odone A. COVID-19 in Italy: impact of containment measures and prevalence estimates of infection in the general population. Acta Biomed. 2020;91:175–9.

12. The Washington Post, Why we should be skeptical of China’s coronavirus quarantine. 2020. https://www.washingtonpost.com/outlook/why-we-should-be-skeptical-of-chinas-coronavirus-quarantine/2020/01/24/51b711ca-3e2d-11ea-8872-5df698785a4e_story.html (28 April 2020, date last accessed).

13. Bloomberg, China’s Unproven Antiviral Solution: Quarantine of 40 Million. 2020. https://www.bloomberg.com/news/articles/2020-01-24/china-s-unproven-antiviral-solution-quarantine-of-40-million (28 April 2020, date last accessed).

14. Pan A, Liu L, Wang C, et al. Association of Public Health Interventions With the Epidemiology of the COVID-19 Outbreak in Wuhan, China. JAMA. 2020;323:1915–23.

15. Gatto M, Bertuzzo E, Mari L, et al. Spread and dynamics of the COVID-19 epidemic in Italy: Effects of emergency containment measures. Proc Natl Acad Sci U S A. 2020;117:10484–91.

16. Wang KW, Gao J, Wang H, et al. Epidemiology of 2019 novel coronavirus in Jiangsu Province, China after wartime control measures: A population-level retrospective study. Travel Med Infect Dis. 2020;35:101654.

17. Lasry A, Kidder D, Hast M, et al. Timing of Community Mitigation and Changes in Reported COVID-19 and Community Mobility - Four U.S. Metropolitan Areas, February 26-April 1, 2020. MMWR Morb Mortal Wkly Rep. 2020;69:451–7.

18. Kraemer MUG, Yang CH, Gutierrez B, et al. The effect of human mobility and control measures on the COVID-19 epidemic in China. Science. 2020.

19. Chinese Center for Disease Control and Prevention. http://2019ncov.chinacdc.cn/2019-nCoV/ (28 April 2020, date last accessed).

20. Baidu Migration. http://qianxi.baidu.com/ (28 April 2020, date last accessed).

21. Feng L, Zhao N, Yao X, et al. Histidine-tryptophan-ketoglutarate solution vs. University of Wisconsin solution for liver transplantation: a systematic review. Liver Transpl. 2007;13:1125–36.

22. Gao XF, Wang L, Liu GJ, et al. Rifampicin plus pyrazinamide versus isoniazid for treating latent tuberculosis infection: a meta-analysis. Int J Tuberc Lung Dis. 2006;10:1080–90.

23. Lai S, Ruktanonchai NW, Zhou L, et al. Effect of non-pharmaceutical interventions to contain COVID-19 in China. Nature. 2020.

24. The State Council, Notice of the State Council on readjustment of the Standards for The Division of Urban Dimensions. 2014 http://www.gov.cn/zhengce/content/2014-11/20/content_9225.htm (28 April 2020, date last accessed).

25. Jia JS, Lu X, Yuan Y, Xu G, Jia J, Christakis NA. Population flow drives spatio-temporal distribution of COVID-19 in China. Nature. 2020;582:389–94.

26. Yen MY, Chiu AW, Schwartz J, et al. From SARS in 2003 to H1N1 in 2009: lessons learned from Taiwan in preparation for the next pandemic. J Hosp Infect. 2014;87:185–93.

27. Chan KP. Control of severe acute respiratory syndrome in singapore. Environ Health Prev Med. 2005;10:255–9.

28. Rashid H, Ridda I, King C, et al. Evidence compendium and advice on social distancing and other related measures for response to an influenza pandemic. Paediatr Respir Rev. 2015;16:119–26.

29. Cauchemez S, Ferguson NM, Wachtel C, et al. Closure of schools during an influenza pandemic. Lancet Infect Dis. 2009;9:473–81.

30. Kawaguchi R, Miyazono M, Noda T, Takayama Y, Sasai Y, Iso H. Influenza (H1N1) 2009 outbreak and school closure, Osaka Prefecture, Japan. Emerg Infect Dis. 2009;15:1685.

31. Ishola DA, Phin N. Could influenza transmission be reduced by restricting mass gatherings? Towards an evidence-based policy framework. J Epidemiol Glob Health. 2011;1:33–60.

32. Zhang C, Chen C, Shen W, et al. Impact of population movement on the spread of 2019-nCoV in China. Emerg Microbes Infect. 2020:1–28.

33. Hanming Fang, Long Wang, Yang Yang. National Bureau of Economic Research working paper series - Human Mobility Restrictions and the Spread of the Novel Coronavirus (2019-NCOV) in China. 2020. https://www.nber.org/papers/w26906.pdf (17 May 2020, date last accessed).

34. Chinazzi M, Davis JT, Ajelli M, et al. The effect of travel restrictions on the spread of the 2019 novel coronavirus (COVID-19) outbreak. Science. 2020;368:395–400.

35. Lau H, Khosrawipour V, Kocbach P, et al. The positive impact of lockdown in Wuhan on containing the COVID-19 outbreak in China. J Travel Med. 2020;27:taaa037.

36. Kasaie P, David Kelton W, Ancona RM, Ward MJ, Froehle CM, Lyons MS. Lessons Learned From the Development and Parameterization of a Computer Simulation Model to Evaluate Task Modification for Health Care Providers. Acad Emerg Med. 2018;25:238–49.

37. Song X, Wang C, Hu H, Huang T, Jin J. A Finite Element Study of the Dynamic Response of Brain Based on Two Parasagittal Slice Models. Comput Math Methods Med. 2015;2015:816405.

38. Kelso JK, Milne GJ, Kelly H. Simulation suggests that rapid activation of social distancing can arrest epidemic development due to a novel strain of influenza. BMC Public Health. 2009;9:117.

39. Nussbaumer-Streit B, Mayr V, Dobrescu AI, et al. Quarantine alone or in combination with other public health measures to control COVID-19: a rapid review. Cochrane Database Syst Rev. 2020;4:CD013574.

40. Wesolowski A, Eagle N, Tatem AJ, et al. Quantifying the impact of human mobility on malaria. Science. 2012;338:267–70.

41. Wesolowski A, Qureshi T, Boni MF, et al. Impact of human mobility on the emergence of dengue epidemics in Pakistan. Proc Natl Acad Sci U S A. 2015;112:11887–92.

42. Buckee CO, Balsari S, Chan J, et al. Aggregated mobility data could help fight COVID-19. Science. 2020;368:145–6.

